# Assessing genetic overlap and causality between blood plasma proteins and Alzheimer’s Disease

**DOI:** 10.1101/2021.04.21.21255751

**Authors:** Alex Handy, Jodie Lord, Rebecca Green, Jin Xu, Dag Aarsland, Latha Velayudhan, Abdul Hye, Richard Dobson, Petroula Proitsi, on behalf of the Alzheimer’s Disease Neuroimaging initiative, Add NeuroMed, and the GERAD1 Consortium

**Author notes:** Correspondence to Petroula Proitsi. Data used in preparation of this article were obtained from the Alzheimer’s Disease Neuroimaging Initiative (ADNI) database (adni.loni.usc.edu). As such, the investigators within the ADNI contributed to the design and implementation of ADNI and/or provided data but did not participate in analysis or writing of this report. A complete listing of ADNI investigators can be found at http://adni.loni.usc.edu/wp-content/uploads/how_to_apply/ADNI_Acknowledgement_List.pdf. The members of Alzheimer’s Disease Neuroimaging Initiative are listed in the Appendix. Data used in the preparation of this article were obtained from the Genetic and Environmental Risk for Alzheimer’s disease (GERAD1) Consortium. As such, the investigators within the GERAD1 consortia contributed to the design and implementation of GERAD1 and/or provided data but did not participate in analysis or writing of this report.

## Abstract

**Background:** Blood plasma proteins are modifiable and have been associated with Alzheimer’s disease (AD), but understanding which proteins are on the causal pathway remains challenging.

**Objective:** Investigate the genetic overlap between candidate proteins and AD using polygenic risk scores (PRS) and interrogate their causal relationship using bi-directional Mendelian Randomization (MR).

**Methods:** Following a literature review, 31 proteins were selected for PRS analysis. PRS were constructed for prioritised proteins with and without the apolipoprotein E region (APOE+/- PRS) and tested for association with AD status across three cohorts (n=6244). An AD PRS was also tested for association with protein levels in one cohort (n=410). Proteins showing association with AD were taken forward for MR.

**Results:** For APOE e3, apolipoprotein B-100, and C-reactive protein (CRP), protein APOE+ PRS were associated with AD below Bonferroni significance (pBonf, p-value <0.00017). No protein APOE-PRS or AD PRS (APOE+/-) passed pBonf. However, vitamin D-binding protein (protein PRS APOE-, p-value=0.009) and insulin-like growth factor-binding protein 2 (AD APOE- PRS p-value=0.025, protein APOE-PRS p-value=0.045) displayed suggestive signals and were selected for MR. In bi-directional MR, none of the 5 proteins demonstrated a causal association (p-value<0.05) in either direction.

**Conclusion:** Apolipoproteins and CRP PRS are associated with AD and provide a genetic signal linked to a specific, modifiable risk factor. Whilst evidence of causality was limited, this study was conducted in a moderate sample size and provides a framework for larger samples with greater statistical power.

## INTRODUCTION

Over 50 million people currently live with dementia worldwide, a figure forecast to rise to 152 million by 2050 as global populations live longer [1]. The most common form of dementia is late-onset Alzheimer’s disease (AD) [2], where individuals suffer severe, progressive cognitive decline and a range of neuropsychiatric symptoms [3] from their mid 60s until death. AD is a highly heritable [4], polygenic trait [5, 6] with a wide range of known genetic and environmental risk factors [7, 8]. However, the precise aetiology of AD remains unexplained [9] and no disease altering treatments exist [10, 11].

Endophenotypes representing traits closer to a hypothesized biological risk factor can help unpack AD aetiology and provide modifiable targets for intervention. For example, a wide range of blood plasma proteins have been associated with AD [12] and provide a potential avenue for disease diagnosis and treatment. Promisingly, prediction of AD diagnosis using plasma levels of amyloid-β (Aβ) [13] and tau [14] is improving towards clinical level. However, measuring known AD protein neuropathological end-products (Aβ and tau) provides limited explanation of how other plasma proteins may mediate AD risk. For example, Kiddle et al’s systematic review identified that 4 proteins (apolipoprotein E (APOE), alpha-2-macroglobulin, complement C3 and alpha-1-antitrypsin) were associated with AD in at least 5 cohorts [12]. APOE and the complement pathway have been consistently implicated in functional and genetic studies of AD risk [15, 16] and recent Mendelian Randomization (MR) studies suggest lower levels of APOE and complement C3 in plasma may be causal for AD [17, 18].

One way to explore the role of plasma proteins further is to assess their genetic overlap with AD. Recent improvements in protein assay technology have enabled the creation of a genetic atlas for plasma proteins [19]. Over 3000 proteins now have publicly available genetic summary statistics [19], allowing polygenic risk scores (PRS) to be constructed for individual proteins. PRS represent aggregate genetic propensity for a trait and so if associated with another trait imply a degree of shared genetics influences both traits. For example, a higher AD PRS has been associated with lower cognitive ability in individuals without dementia [20] and with increased levels of the promising AD biomarker p-tau181[21]. If a protein PRS is associated with AD, this provides a genetic signal linked to a specific, modifiable biological risk factor, something which remains a challenge for genome wide association studies (GWAS) [22]. PRS can also be calculated for individuals meaning protein PRS associated with AD could inform AD diagnosis prediction.

However, testing the association of PRS with a trait does not demonstrate causality. For example, a plasma protein PRS associated with AD may simply indicate shared genetic variants which effect traits or pathways unrelated to disease pathogenesis [23]. MR provides a method to test whether an exposure causally effects an outcome by using genetic variants as instrumental variables in a construct similar to a randomized control trial. This works because an individual’s genes are effectively randomised at birth enabling the creation of a quasi-intervention group who have a genetic disposition for an exposure [24]. In AD, MR has demonstrated the protective effect of higher cognitive ability and educational attainment in two large scale studies [7, 25] and has indicated several blood metabolites are on the causal disease pathway[26]. For plasma proteins specifically, MR has primarily been deployed as part of large scale non-targeted, phenome-wide MR analysis [27, 28]. For example, Zheng et al identified 111 causal associations between 65 proteins and 52 disease-related phenotypes (p<3.5×10^−7^) including sialic acid binding Ig-like lectin 3 (CD33) with AD, supporting previous GWAS results [29, 30]. Whilst phenome-wide MR designs excel at providing a multi-trait matrix of potential causal signals, they lack the flexibility to unpack disease specific relationships. For example, 61 out of 62 proteins associated with AD at a p-value less than 0.05 in Zheng et al were tested using the Wald Ratio with only 1 SNP as an instrumental variable [27]. Disease specific approaches have more flexibility to relax assumptions which can help increase statistical power and enable more robust statistical sensitivity analyses [23].

The primary objective of this study was to explore the genetic overlap between AD and plasma proteins using PRS and to assess whether individual plasma proteins play a causal role in AD aetiology using MR (see *Figure 1* for illustrative overview of study design). This study identified a shortlist of plasma proteins from existing literature that have been robustly associated with AD or AD endophenotypes and have publicly available genetic summary data. PRS models were then created for each shortlisted protein and tested for association with AD across three consortium cohorts, Genetic and Environmental Risk in Alzheimer’s Disease (GERAD1), Alzheimer’s Disease Neuroimaging Initiative (ADNI) and AddNeuroMed (ANM). An AD PRS was also constructed with publicly available genetic summary data from the largest case ascertained AD GWAS to date [8] and used to test each protein for bi-directional association in ANM where individual level plasma protein data was available. Lastly, for plasma proteins with PRS that demonstrated significant associations with AD (see Materials and Methods) in one or both PRS analyses, two sample bi-directional MR was conducted to test for causality (Figure 1).

**Figure 1:**
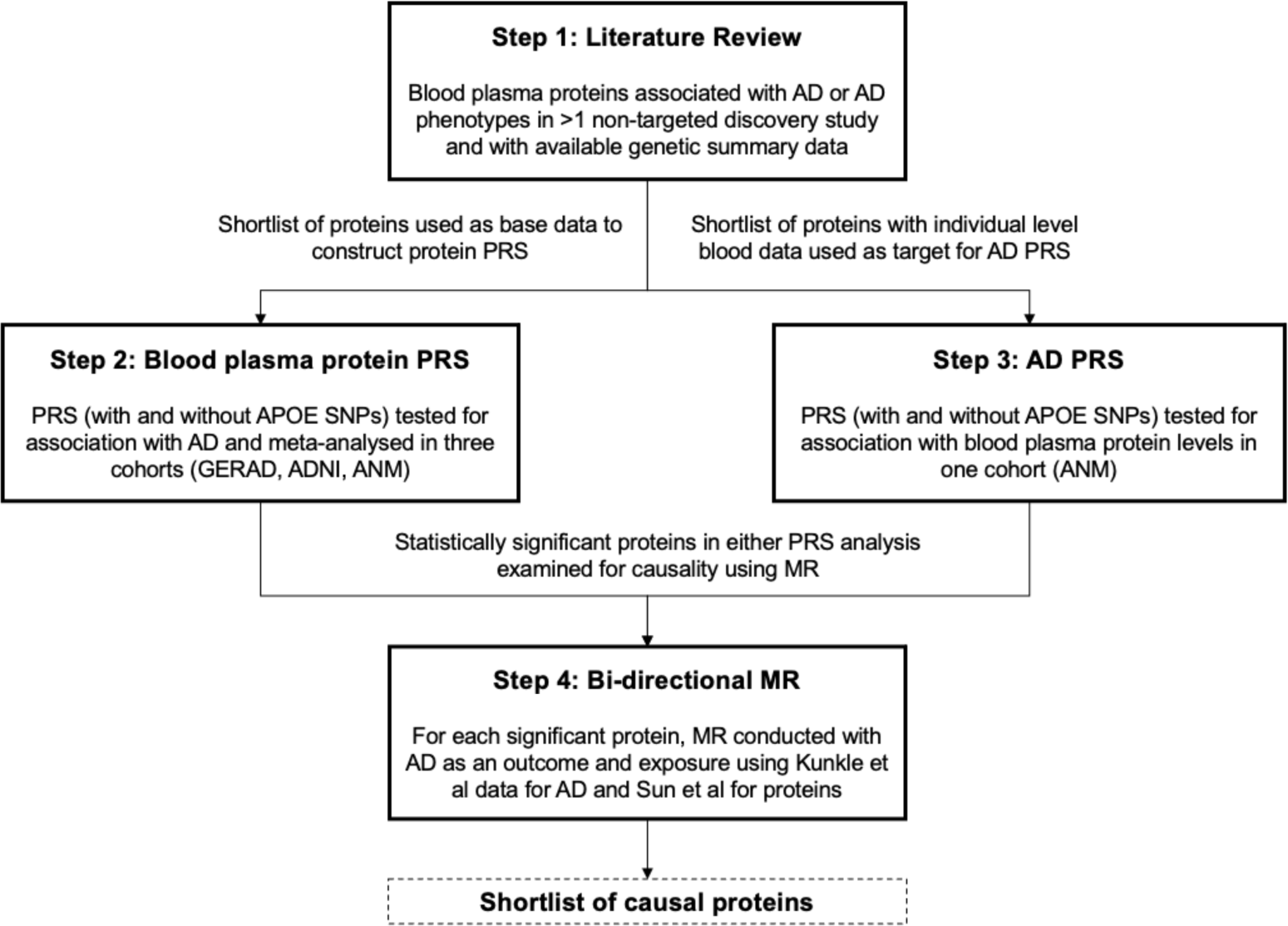
Illustrative overview of study design outlining the four key steps in the study workflow – literature review, plasma protein PRS, AD PRS and bi-directional MR.

## MATERIALS AND METHODS

### Plasma protein candidates

A shortlist of plasma proteins was compiled for analysis from previous AD discovery studies to represent known protein candidates for genetic exploration. A panel of 163 proteins previously associated with AD or AD related phenotypes from a systematic review of 21 non-targeted human AD blood protein discovery studies up to 2014 was used as the baseline [12]. An additional literature review was then conducted to cover the period from 2014-2019 and applied the same screening criteria (non-targeted discovery studies, except for panel based studies with >100 candidates) [12] to generate the candidate list (*further details in Supplementary Notes*). Proteins were then selected for the shortlist if they passed two criteria. Firstly, the association replicated in >1 study and, secondly, genetic summary data was available for the protein from Sun et al’s human plasma protein GWAS [19]. At the time of analysis, Sun et al was the largest, plasma protein GWAS using SomaLogic with publicly available data (downloaded and cross referenced from http://www.phpc.cam.ac.uk/ceu/proteins/).

### Plasma protein data preparation

Plasma protein genetic summary data was acquired from Sun et al’s GWAS of 3622 proteins in 3301 healthy participants from the INTERVAL study (*further details in Supplementary Notes*), a randomized trial of 45,000 blood donors across 25 centres in England [31]. Summary association results were made available on http://www.phpc.cam.ac.uk/ceu/proteins/ and were downloaded for proteins on the shortlist. Files were then uploaded to Rosalind, King’s College London’s high-performance computing facility, for further quality control (QC) and analysis. Further QC was conducted using R.3.6.0 and was based primarily on aligning to the protocol recommended for PRS analysis [32]. Specifically, remaining duplicates, non bi-allelic and non-target data overlapping variants were removed and SNP rsIDs were added and aligned with the target dataset (see https://github.com/AlexHandy1/ad-genetic-overlap-analysis for analysis scripts). Observed SNP heritability (h^2 SNP^) and cross protein genetic correlation (rg) was estimated for each protein using linkage disequilibrium score regression (LDSR) with the Python LDSC package (Version 1.0.0) sourced from https://github.com/bulik/ldsc.

### AD data preparation

Individual level genotype data for AD cases and controls was acquired from three consortium studies, GERAD1, ADNI and ANM.

GERAD1 is a European consortium (https://gtr.ukri.org/projects?ref=G0902227)[33]. The GERAD1 sample comprised up to 3292 AD cases and 1223 controls. Genetic data from a subset of 4515 participants was made available for this analysis prior to QC *(further details in Supplementary Notes)*.

ADNI is a longitudinal, multi-centre North American study initiated in 2004 [34] and now in its fourth wave (ADNI1, ADNI-GO, ADNI2 and ADNI3) collecting clinical, imaging, genetic and biomedical biomarkers for AD. Genetic data from 1674 participants (not including ADNI3) was made available for this analysis prior to QC (*further details in Supplementary Notes*).

The ANM is a European consortium, initiated in 2008 with the aim to establish biomarkers for AD [35]. The Dementia Case Register (DCR) is a follow-up of ANM, with UK subjects recruited from the Maudsley and King’s Healthcare Partners Dementia Case Register [36]. Genetic data from 1063 participants from ANM (including DCR) was made available for this analysis prior to QC (*further details in Supplementary Notes*). In addition to full clinical and demographic data, 410 participants had plasma protein data. Plasma protein data was collected using SomaLogic’s multiplexed, aptamer-based assay (SOMAscan) with SOMAmers for 1016 proteins. Normalised, log2 transformed residuals were used as the phenotype after regression with age, sex, batch and 10 principal components.

For all three consortium datasets, standard genetic QC procedures were applied (removal of non-autosomal chromosomes, alleles with minor allele frequency <1%, genotypes with call rate <98% and Hardy Weinberg deviations at 1×10^-5^) based on protocol by Coleman [37].

### Plasma protein PRS

A PRS was constructed with the post QC Sun et al GWAS data (“base data”) for each shortlist protein using PRSice-2 software (version 2.3.1e)[38]. PRSice automatically removes strand ambiguous SNPs and SNPs that are not present in both base and target data. SNPs were further “clumped” if LD (measured in r^2^) was >0.1 within a window of 250 kilobases with the SNP with the lowest GWAS p-value within each region retained. After clumping, PRS were calculated with SNPs under 10 pre-defined p-value thresholds (5e-08, 5e-05, 5e-04, 0.0001, 0.001, 0.01, 0.05, 0.1, 0.2, 0.5, 1). PRS at all 10 thresholds for each protein were then independently tested for association with each individual AD dataset (GERAD1, ADNI and ANM) using logistic regression. Age, sex and 7 principal components to control for population stratification were included as covariates to create a null model (covariates only), which was subtracted from the full model (covariates and PRS), to provide a Lee adjusted r^2^ [39] assuming an AD prevalence of 7% [40].

Protein PRS, for each selected protein, were tested for association with AD with the APOE region (992 SNPs within 750 kilobases of rs429358 on chromosome 19) included (APOE+) and excluded (APOE-) for all participants. APOE SNPs were removed to test whether protein PRS associations held without the known, strong effects of APOE alleles on AD risk [41]. As a secondary analysis to explore sex and age specific associations, PRS were constructed and tested for subgroups stratified by male, female and 70 years and over (>=70) to test for gender and age specific effects. Lastly, random-effects meta-analysis was conducted on the PRS results at each p-value threshold across the three AD datasets using R and a restricted maximum likelihood (REML) estimator from the metafor R package [42]. Meta-analysed results were ranked by lowest p-value to identify the most significant PRS model threshold for each protein-AD association. A Bonferroni corrected p-value of 0.00017 (0.05 / (number of proteins x number of PRS p-value thresholds tested)) was estimated and used as the threshold for significance.

### AD PRS

An AD PRS was constructed using the meta-analysed stage 1 discovery results from Kunkle et al’s AD GWAS [8] of 21,982 AD cases and 41,944 cognitively normal controls (“base data”) downloaded from https://www.niagads.org/datasets/ng00075. Kunkle et al was selected to provide statistical power and a clinical phenotype as the largest case ascertained AD GWAS to date. QC was applied to the base data of 11,480,632 variants to remove NA’s, variants without an rsID and non bi-allelic variants leaving 10,528,610 variants for PRS analysis. As rsIDs were already provided in the AD base data and aligned to GRCh37, non-target data overlapping variants were removed with PRSice during the PRS analysis. P-value thresholds and clumping configuration settings in PRSice were kept constant with the protein PRS analysis. No covariates were included as age, sex, batch and population stratification were already controlled for in the production of the protein residuals phenotype which was used as the target data. AD PRS were tested for association with each individual shortlist protein using linear regression and the PRS model with the best fit (measured by lowest p-value) was presented for each protein. As with the protein PRS, AD PRS were tested for association with and without APOE SNPs and secondary analysis was conducted on subsets stratified by gender (male, female) and age (>=70). Bonferroni correction was estimated and applied to control for multiple testing as in the protein PRS analysis (with number of tests adjusted for the number of proteins with individual level blood data available for analysis).

### Bi-directional MR

MR analysis was performed using the MR Base R package [43] on a subset of proteins from the PRS analyses. Proteins were for selected for MR if they had a p-value below Bonferroni significance in either PRS analysis (protein PRS to AD APOE+/- or AD PRS to protein APOE+/-) or if they were nominally significant (p-value < 0.05) in both directions (protein PRS to AD APOE+/- and AD PRS to protein APOE+/-). Given Bonferroni is a conservative threshold, if no protein passed Bonferroni significance, the strongest protein association with a p-value below 0.05 was also considered. Univariate MR was performed with each protein as the exposure and AD as the outcome. Genetic instrument SNPs for each protein were selected from Sun et al at two p-value thresholds for analysis (5×10^-8^ and 5×10^-6^).

The less stringent 5×10^-6^ threshold was applied to ensure SNPs were available for all proteins with the noted limitation of introducing potential weak instrument bias. Selected SNPs were then clumped within a 250kb window at LD r^2^ < 0.001. F statistics were generated for each SNP (SNP-exposure effect size^2 / SNP-exposure standard error^2) to test for weak instrument bias and excluded if < 10 [44]. The remaining SNPs were further pruned if they were associated with any of the other proteins or with AD directly (p-value < 5×10^-8^ in Kunkle et al GWAS). This was implemented to exclude SNPs that may affect AD through a pathway other than the exposure protein (horizontal pleiotropy) [45]. SNPs were also removed in the APOE region (chromosome 19, base-pairs 4500000-4580000) as potential confounders that violate MR’s core assumptions, given their known association with AD [46]. Protein exposure SNPs and AD outcome SNPs were harmonized and tested with alleles assumed to be on the forward strand (no palindromic SNPs removed). As a secondary test, the MR analysis was also run with palindromic SNPs flipped and removed if non-inferable. Causal estimates were estimated using inverse variance weighted (IVW) two sample MR and sensitivities were tested with MR-Egger, weighted median and leave one out analysis (*further details in Supplementary Notes*). To test for causality in the opposite direction, this analysis pipeline was repeated with AD as the exposure (using AD SNPs with p-value < 5×10^-8^ from Kunkle et al stages 1, 2 and 3) and each protein as the outcome.

An interactive web dashboard was built with R Shiny to present the full PRS and MR results (available at https://alexhandy1.shinyapps.io/ad-genetic-overlap-web-results/) with the key results reported herein.

## RESULTS

### DATA PREPARATION

The literature review provided 4 new studies [36,47–49] adding 14 new proteins and bringing the total candidate protein list to 175. From the 175 candidate proteins, 31 passed the shortlist inclusion criteria (>1 study replication, GWAS data available, see *Table 1*).

**Table 1:** Protein shortlist for analysis ordered by number of studies replicated in from literature review.

| Protein name | UniProt ID | Studies replicated in (N) | SOMAmer ID (Sun et al study) |
| --- | --- | --- | --- |
| Pancreatic prohormone | P01298 | 7 | PPY.4588.1.2 |
| Apolipoprotein E (isoform ε3) | P02649 | 6 | APOE.2937.10.2 |
| Complement factor H | P08603 | 5 | CFH.4159.130.1 |
| Plasma protease C1 inhibitor | P05155 | 5 | SERPING1.4479.14.2 |
| Complement C3 | P01024 | 5 | C3.2755.8.2 |
| Fibrinogen (D-dimer) | P02671, P02675, P02679 | 5 | FGA.FGB.FGG.4907.56.1 |
| Serum amyloid P-component | P02743 | 4 | APCS.2474.54.5 |
| Haptoglobin | P00738 | 3 | HP.3054.3.2 |
| Interleukin-3 | P08700 | 3 | IL3.4717.55.2 |
| Complement C4-A/B | P0C0L4 P0C0L5 | 3 | C4A.C4B.4481.34.2 |
| Interleukin-10 | P22301 | 3 | IL10.2773.50.2 |
| Vitronectin | P04004 | 3 | VTN.13125.45.3 |
| Insulin-like growth factor-binding protein 2 | P18065 | 3 | IGFBP2.2570.72.5 |
| Angiopoietin-2 | O15123 | 2 | ANGPT2.13660.76.3 |
| Apolipoprotein B-100 | P04114 | 2 | APOB.2797.56.2 |
| C-C motif chemokine 26 | Q9Y258 | 2 | CCL26.9168.31.3 |
| C-reactive protein | P02741 | 2 | CRP.4337.49.2 |
| Clusterin | P10909 | 2 | CLU.4542.24.2 |
| Granulocyte colony-stimulating factor | P09919 | 2 | CSF3.8952.65.3 |
| Interleukin-13 | P35225 | 2 | IL13.3072.4.2 |
| Interleukin-8 | P10145 | 2 | CXCL8.3447.64.2 |
| Kit ligand | P21583 | 2 | KITLG.9377.25.3 |
| Matrix metalloproteinase-9 | P14780 | 2 | MMP9.2579.17.5 |
| Natriuretic peptides B | P16860 | 2 | NPPB.3723.1.2 |
| Plasminogen | P00747 | 2 | PLG.3710.49.2 |
| Resistin | Q9HD89 | 2 | RETN.3046.31.1 |
| Serotransferrin | P02787 | 2 | TF.4162.54.2 |
| Tenascin | P24821 | 2 | TNC.4155.3.2 |
| Tumor necrosis factor | P01375 | 2 | TNF.5936.53.3 |
| Vascular cell adhesion protein 1 | P19320 | 2 | VCAM1.2967.8.1 |
| Vitamin D-binding protein | P02774 | 2 | GC.6581.50.3 |

Sun et al’s GWAS provided summary statistics for 3301 participants (see characteristics in *Supplementary Table 1*) covering 10,572,788 variants for each protein, with 5,210,103 variants included for PRS analysis after additional QC for this study (see *Supplementary Table 2*). Average h^2 SNP^ across the proteins was 0.10, however, results were treated as indicative given the average standard error was 0.16 (including 8 proteins with h^2 SNP^ less than 0) (see Supplementary Notes for further details).

For the AD cohort, 6244 participants were available for analysis from GERAD1, ADNI and ANM (see *Table 2*) with 5,218,413 overlapping variants included for PRS analysis (see Supplementary Notes for further details).

**Table 2:** Summary characteristics of GERAD1, ADNI and ANM participants post QC.

|  | **Total** | **AD**<br>**Cases** | **Controls** | **Males** | **Females** | **70 and**<br>**over** | **Mean**<br>**age** |
| --- | --- | --- | --- | --- | --- | --- | --- |
| **GERAD1**<br>**(N)** | 4492 | 3277 | 1215 | 1640 | 2852 | 3189 | 70.5 |
| **ADNI (N)** | 1007 | 639 | 368 | 559 | 448 | 886 | 78.2 |
| **ANM (N)** | 745 | 371 | 374 | 309 | 436 | 641 | 78.2 |
| **Total (N)** | **6244** | **4287** | **1957** | **2508** | **3736** | **4716** |  |

For ANM, 410 participants with plasma protein data remained for analysis after QC (see *Table 3*).

**Table 3:**
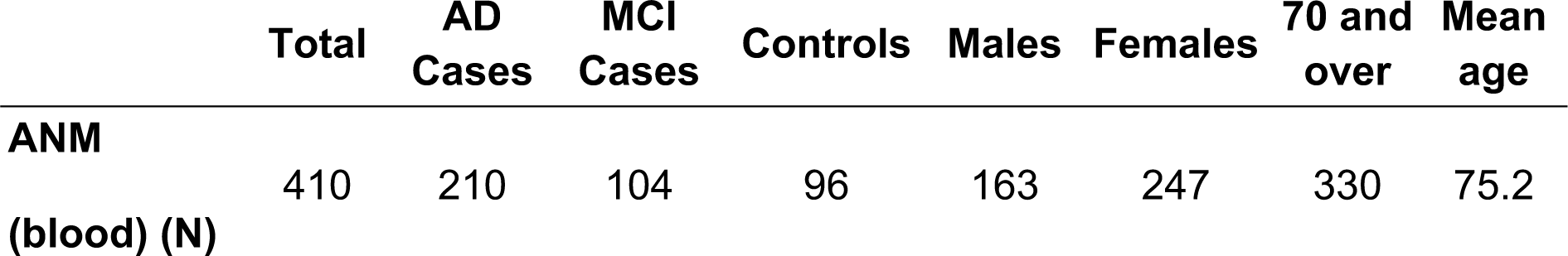
Summary characteristics of ANM participants with plasma protein data post QC.

## PRS RESULTS

### Plasma protein PRS

In all participants, APOE+ PRS for APOE ε3 (p-value = 6.5×10^-21^), apolipoprotein B-100 (APOB-100, p-value = 6.7×10^-20^) and C-reactive protein (CRP, p-value = 1.5×10^-8^) were associated with AD at Bonferroni significance (p-value < 0.00017) (see *Table 4 and full results* at https://alexhandy1.shinyapps.io/ad-genetic-overlap-web-results/). No other protein APOE+ PRS passed Bonferroni significance in all participants or subgroups. For APOE-PRS, no proteins, including APOE, APOB and CRP, passed Bonferroni significance in all participants or subgroups. In all participants, APOE-PRS for vitamin D-binding protein (VDBP) presented the strongest signal (p-value = 0.009) with 7 other proteins passing nominal significance (p-value < 0.05). The 3 proteins passing Bonferroni significance in APOE+ PRS (APOE ε3, APOB-100 and CRP) and the strongest APOE-PRS signal in all participants (VDBP) were deemed signals warranting further exploration in MR analysis.

**Table 4:** Summary of proteins with APOE+ PRS associations from meta-analysed logistic regression with AD in all participants below Bonferroni significance.

| Protein | PRS p-value threshold* | SNPs in PRS (N) | Beta** | 95% CI Lower | 95% CI Upper | R<sup>2</sup>*** | P-value |
| --- | --- | --- | --- | --- | --- | --- | --- |
| APOE ε3 | 5.0E-08 | 5 | 0.26 | 0.20 | 0.31 | 0.01 | 6.5E-21 |
| APOB-100 | 5.0E-08 | 2 | 0.58 | 0.46 | 0.70 | 0.01 | 6.7E-20 |
| CRP | 5.0E-08 | 5 | -0.31 | -0.42 | -0.20 | 0.01 | 1.5E-08 |

\* P-value from protein GWAS below which SNPs were included in PRS. PRS p-value threshold for most significant PRS model (based on meta-analysed p-value) presented.
\*\* Estimated based on normalised, per standard deviation of PRS
\*\*\* Estimated from meta-analysis outputs using the formula $R^2 = \left(\frac{z}{\sqrt{n-2+z^2}}\right)^2$ where z is the Z-score for the protein PRS and n is the total sample size.

### AD PRS

AD PRS models were tested for association with 26 of the 31 shortlist proteins in 1 ANM cohort (due to data availability) and in the same 3 subgroups (males, females and >=70). In all participants and subgroups, no proteins were associated with AD APOE+ or APOE-PRS at Bonferroni significance (p-value < 0.00019, view full results at https://alexhandy1.shinyapps.io/ad-genetic-overlap-web-results/). In all participants, Haptoglobin presented the strongest association with AD APOE+ PRS (p-value = 0.0107), with CRP (p-value = 0.0108) the other protein to pass nominal significance. For AD APOE-PRS, Complement factor H was the strongest signal (p-value = 0.021) with 6 other proteins passing nominal significance. Insulin-like growth factor-binding protein 2 (IGFBP2) was one of these proteins (p-value = 0.025) and came close to Bonferroni significance in >=70 (p-value = 0.00026). Given IGFBP2 was also the only protein to display a nominally significant association for APOE-PRS in both directions (AD APOE-PRS p-value = 0.025, protein APOE-PRS p-value = 0.045) it was selected for MR analysis.

### BI-DIRECTIONAL MR RESULTS

MR analysis was conducted with 5 proteins, APOE ε3, APOB-100, CRP, IGFBP2 and VDBP to test for casual associations. Only 2 proteins (CRP and VDBP) had valid SNP instruments at 5×10^-8^ but all proteins had available SNP instruments at 5×10^-6^. No proteins passed nominal significance (p-value < 0.05) using IVW with SNPs selected at 5×10^-8^ or 5×10^-6^ (see *Figure 2* and view full results at https://alexhandy1.shinyapps.io/ad-genetic-overlap-web-results/). Overall, exclusion of non-inferable palindromic SNPs produced similar results across all methods. All instruments selected had an F statistic > 10 and there was limited evidence of horizontal pleiotropy (no Egger intercept p-value < 0.05) or heterogeneity between SNPs (no Cochran’s Q p-value < 0.05 for protein across multiple methods). There was also no evidence of reverse causality when AD was tested as an exposure.

**Figure 2:**
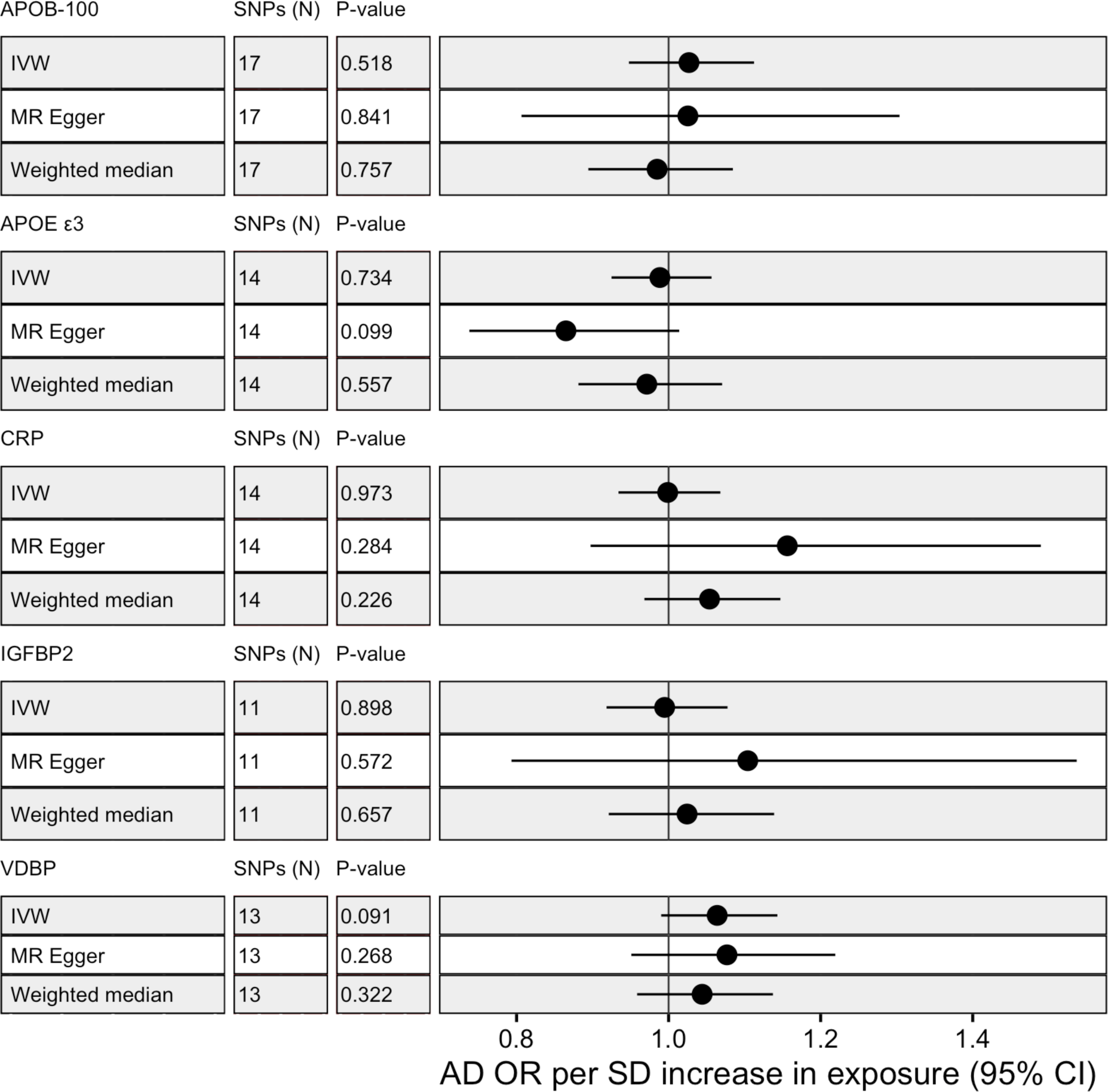
MR results with protein as exposure and AD as outcome for IVM, MR-Egger and weighted median with alleles assumed on forward strand for harmonisation with SNP instruments selected at 5×10^-6^.

## DISCUSSION

### Main Findings

This study set out to identify if a shortlist of plasma proteins genetically overlap with AD by testing if PRS for these proteins were associated with AD. For proteins with evidence of genetic overlap, MR was conducted to test whether exposure to the protein was causal. The findings provide strong evidence that APOE ε3, APOB-100 and CRP genetically overlap with AD and, therefore, identifies a genetic signal linked to a set of specific, modifiable risk factors. Moreover, this overlap appears to be driven by the APOE genotype, providing evidence that APOE’s strong genetic effect on AD [41] may be partially linked to plasma proteins. Apolipoproteins in plasma have been linked to AD risk through their role in regulating cholesterol [50], with increased total cholesterol (TC) associated with higher risk [51, 52] whilst increased high-density lipoprotein cholesterol (HDL-C) appears protective [53]. CRP has also been suggested as a causal factor in AD through its pro-inflammatory role in the immune response [54, 55]. Observational evidence is nuanced, with increased levels of CRP in midlife associated with multiple dementias in later life [56] contrasting with lower levels of CRP found in individuals who actively have AD [57].

Despite the evidence of genetic overlap and supporting observational evidence, our MR analysis found no evidence of causality for APOE ε3, APOB-100 or CRP. This aligns with larger MR studies that found no causal associations between AD and CRP [25], TC or HDL-C [58, 59], the hypothesized mechanisms of action for apolipoproteins. However, a more recent MR meta-analysis found that both TC and HDL-C showed causal associations with AD and demonstrated a dose response relationship linked to APOE genotype [60]. This also aligns with Rasmussen et al’s larger MR study which suggests APOE in plasma is causal for AD[18]. Unfortunately due to data availability, our study was only able to measure APOE isoform ε3 rather than the higher effect isoforms ε2 and ε4 [61] suggesting our null MR result may be due to isoform rather than a lack of causality.

Independently of APOE, VDBP and IGFBP2 presented suggestive signals of genetic overlap with AD in our study. Research investigating the effect of VDBP and IGFBP2 on AD is less well developed. VDBP has been shown to attenuate Aβ aggregation in vitro and in mice [62] and a recent MR study identified a causal relationship between increased levels VDBP and reduced AD risk [63]. However, Zhang et al used only 1 SNP as an instrumental variable and were not able to identify a causal association with other AD phenotypes. IGFBP2 has been shown to restrict the neuroprotective effects of insulin growth factors [64] and increased levels have been associated with higher AD risk and lower cognitive performance [65]. In our MR analysis, neither VDBP or IGFBP2 demonstrated robust evidence of causality. To the best of our knowledge, this is the first study to conduct a targeted MR on IGFBP2 and AD, suggesting further research is required before making a strong inference on causality.

### Limitations and future directions

Our study has three key limitations. Firstly, the initial proteins considered for analysis had to demonstrate prior association with AD and, therefore, represent only a subset of proteins in plasma. Whilst a targeted design was deliberate to lower the risk of false positives and support interpretability, it does increase the probability that causal proteins for AD were not included in the analysis. This exclusion risk was amplified by data availability constraints and a reliance on the SomaLogic platform, with only 53% of candidate proteins from the literature review possessing publicly available genetic summary data at the time of analysis. Most notably, alpha-2-macroglobulin and alpha-1-antitrypsin were each replicated in 6 studies in the literature review (the same number of replications as APOE) but were excluded due to a lack of genetic data. Moving forward, a planned outreach effort to aggregate summary data from other research groups and assay platforms (as demonstrated by [27]) supported by integrating new data [66] could improve protein coverage and increase statistical power.

Secondly, the statistical power to correctly identify true positives was limited across multiple steps of the analysis. Studies are now routinely conducted with samples of 100,000+ individuals for common human traits [67–69] in order to capture the small effect sizes of individual SNPs [70]. Sample size is particularly important for traits with low to moderate heritability where the effect size of individual SNPs is likely to be lower [71]. In this study, estimates of observed SNP heritability (h^2 SNP^) for proteins were themselves hampered by sample size (see https://github.com/bulik/ldsc/wiki/FAQ), but a median h^2 SNP^ of 0.09 (interquartile range 0.006 – 0.22) suggests analysed proteins were at most, moderately heritable. Moderate heritability weakens PRS and MR [72], given PRS and MR instrumental variables are a composite of genetic liability which is dependent on the heritability of a trait. This may partly explain the limited genetic overlap and low variance explained (R^2^) of AD by individual protein PRS (see https://alexhandy1.shinyapps.io/ad-genetic-overlap-web-results/). Low R^2^ may also reflect that proteins often exert their effects as part of large molecular networks rather than as individual entities [73]. Therefore, larger sample sizes and multivariate statistical tests are required to robustly corroborate the lack of causal protein-AD associations in this study.

Lastly, the nature of the phenotype data collected possesses several inherent limitations. Both protein and AD case target data were collected at a single point in time. For AD this means a proportion of controls may have developed into cases and for proteins it means observed variability over time [74, 75] is not captured. For AD there is the added limitation of using a clinical diagnosis as a phenotype. Clinical manifestation is heterogenous [76, 77] and in-life diagnosis can be inaccurate [78, 79] implying that adding endophenotype measures closer to causal biology (e.g. hippocampal volume) and symptomology (e.g. cognitive performance) may allow more precise association analysis.

Future analyses should, therefore, aim to expand protein coverage, increase sample sizes and expand PRS and MR analyses to AD endophenotypes.

## Conclusion

This study provides evidence that apolipoproteins and CRP PRS are associated with AD and identifies a genetic signal linked to a specific, modifiable risk factor. However, none of the proteins tested in MR demonstrated evidence of causality. This study was conducted in a moderate sample size and may have lacked the statistical power to identify true causal associations. Therefore, this study provides a framework for future research to robustly interrogate protein causality in larger samples.

## Data Availability

Data used in preparation of this article was obtained from the Genetic and Environmental Risk for Alzheimer’s disease (GERAD1) Consortium, the Alzheimer’s Disease Neuroimaging Initiative (ADNI) database and the AddNeuroMed consortium.
Access to this data requires an application to this consortia.

https://alexhandy1.shinyapps.io/ad-genetic-overlap-web-results/

## Data Availability

https://alexhandy1.shinyapps.io/ad-genetic-overlap-web-results/

## ACKNOWLEDGEMENTS

This work was also made possible only through generous funding from key funding bodies. Petroula Proitsi is funded by Alzheimer’s Research UK. Jodie Lord is funded by the van Geest endowment fund.

This study represents independent research additionally funded by the National Institute for Health Research (NIHR) Biomedical Research Centre at South London and Maudsley NHS Foundation Trust and King’s College London. The views expressed are those of the author(s) and not necessarily those of the NHS, the NIHR or the Department of Health and Social Care.

The authors acknowledge use of the research computing facility at King’s College London, Rosalind (https://rosalind.kcl.ac.uk), which is delivered in partnership with the National Institute for Health Research (NIHR) Biomedical Research Centres at South London & Maudsley and Guy’s & St. Thomas’ NHS Foundation Trusts, and part-funded by capital equipment grants from the Maudsley Charity (award 980) and Guy’s & St. Thomas’ Charity (TR130505). The views expressed are those of the author(s) and not necessarily those of the NHS, the NIHR, King’s College London, or the Department of Health and Social Care.

GERAD1 Acknowledgements: Cardiff University was supported by the Wellcome Trust, Medical Research Council (MRC), Alzheimer’s Research UK (ARUK) and the Welsh Assembly Government. Cambridge University and Kings College London acknowledge support from the MRC. ARUK supported sample collections at the South West Dementia Bank and the Universities of Nottingham, Manchester and Belfast. The Belfast group acknowledges support from the Alzheimer’s Society, Ulster Garden Villages, N.Ireland R&D Office and the Royal College of Physicians/Dunhill Medical Trust. The MRC and Mercer’s Institute for Research on Ageing supported the Trinity College group. The South West Dementia Brain Bank acknowledges support from Bristol Research into Alzheimer’s and Care of the Elderly. The Charles Wolfson Charitable Trust supported the OPTIMA group.

Washington University was funded by NIH grants, Barnes Jewish Foundation and the Charles and Joanne Knight Alzheimer’s Research Initiative. Patient recruitment for the MRC Prion Unit/UCL Department of Neurodegenerative Disease collection was supported by the UCLH/UCL Biomedical Centre and NIHR Queen Square Dementia Biomedical Research Unit. LASER-AD was funded by Lundbeck SA. The Bonn group was supported by the German Federal Ministry of Education and Research (BMBF), Competence Network Dementia and Competence Network Degenerative Dementia, and by the Alfried Krupp von Bohlen und Halbach-Stiftung. The GERAD1 Consortium also used samples ascertained by the NIMH AD Genetics Initiative.

A proportion of data collection and sharing for this project was also funded by the Alzheimer’s Disease Neuroimaging Initiative (ADNI) (National Institutes of Health Grant U01 AG024904). ADNI is funded by the National Institute on Aging, the National Institute of Biomedical Imaging and Bioengineering, and through generous contributions from the following: Abbott; Alzheimer’s Association; Alzheimer’s Drug Discovery Foundation; Amorfix Life Sciences Ltd.; AstraZeneca; Bayer HealthCare; BioClinica, Inc.; Biogen Idec Inc.; Bristol-Myers Squibb Company; Eisai Inc.; Elan Pharmaceuticals Inc.; Eli Lilly and Company; F. Hoffmann-La Roche Ltd and its affiliated company Genentech, Inc.; GE Healthcare; Innogenetics, N.V.; Janssen Alzheimer Immunotherapy Research & Development, LLC.; Johnson & Johnson Pharmaceutical Research & Development LLC.; Medpace, Inc.; Merck & Co., Inc.; Meso Scale Diagnostics, LLC.; Novartis Pharmaceuticals Corporation; Pfizer Inc.; Servier; Synarc Inc.; and Takeda Pharmaceutical Company. The Canadian Institutes of Health Research is providing funds to support ADNI clinical sites in Canada. Private sector contributions are facilitated by the Foundation for the National Institutes of Health (www.fnih.org). The grantee organization is the Northern California Institute for Research and Education, and the study is coordinated by the Alzheimer’s Disease Cooperative Study at the University of California, San Diego. ADNI data are disseminated by the Laboratory of Neuro Imaging at the University of California, Los Angeles.

## SUPPLEMENTARY MATERIAL

### Supplementary Notes

Additional comments on materials and methods

### Plasma protein shortlist extended literature review

Studies were sourced based on expert recommendation (Proitsi and Hye) and searches on PubMed and Google Scholar for “Alzheimer blood protein discovery”. The search term “Alzheimer blood protein discovery” provided no additional studies passing Kiddle’s screening criteria (non-targeted discovery studies, except for panel based studies with >100 candidates). Therefore, search terms were expanded to include “Alzheimer’s Disease blood plasma proteins”, “Alzheimer’s Disease blood biomarkers” and “Alzheimer’s Disease proteomics”. Proteins were added to the baseline list if individually associated with AD status (at author defined significance threshold, of at least p-value <0.05) or an AD-related phenotype (as defined by list of “outcome variables” in Kiddle et al). AD status associations were prioritised in studies where association was tested with multiple AD-related phenotypes.

### Plasma protein data preparation - Sun et al GWAS

Blood samples were collected in 6-ml EDTA tubes using standard venepuncture protocols and stored at −80 °C before analysis (see [80] for full protocol). Plasma proteins were measured using SomaLogic’s multiplexed, aptamer-based assay (SOMAscan) with 4,034 modified single-stranded DNA SOMAmers that bind to specific protein targets which are then quantified using a DNA microarray (see [19] for further detail). QC procedures resulted in 3283 SOMAmers (mapping to 2994 unique proteins using UniProt identifiers) for GWAS. Individuals were genotyped for 830,000 variants on the Affymetrix Axiom UK Biobank array and imputation was conducted using a combined 1000 Genomes Phase 3-UK10K reference panel via the Sanger Imputation Server. After QC to exclude sex mismatches, low call rates, duplicates, extreme heterozygosity, relatedness and population stratification (see [19] for full protocol), 10,572,788 variants aligned to Genome Reference Consortium genome build 37 (GRCh37) remained for GWAS. Association analysis was performed on the rank-inverse normalised residuals from the linear regression of natural log-transformed protein levels adjusted for age, sex, duration between blood draw and processing (binary, ≤1 day/>1day) and the first three principal components of ancestry from multi-dimensional scaling.

### AD data preparation – GERAD1, ADNI and ANM

GERAD1 participants were genotyped at the Sanger Institute on the Illumina 610-quad chip. These samples were recruited by the Medical Research Council (MRC) Genetic Resource for AD (Cardiff University; Kings College London; Cambridge University; Trinity College Dublin), the Alzheimer’s Research UK (ARUK) Collaboration (University of Nottingham; University of Manchester; University of Southampton; University of Bristol; Queen’s University Belfast; the Oxford Project to Investigate Memory and Ageing (OPTIMA), Oxford University); Washington University, St Louis, United States; MRC PRION Unit, University College London; London and the South East Region AD project (LASER-AD), University College London; Competence Network of Dementia (CND) and Department of Psychiatry, University of Bonn, Germany and the National Institute of Mental Health (NIMH) AD Genetics Initiative. All AD cases met criteria for either probable (NINCDS-ADRDA, DSM-IV) or definite (CERAD) AD. All elderly controls were screened for dementia using the MMSE or ADAS-cog, were determined to be free from dementia at neuropathological examination or had a Braak score of 2.5 or lower. Genotype data is available for ADNI1 and ADN2, typed across three separate genotype chips: ADNI1 (Illumina Human610-Quad BeadChip), ADNI2 (Illumina HumanOmniExpress BeadChip), and OMNI (a combination of phase 1 and 2 participants typed on a high coverage Illumina chip - Omni 2.5M). ADNI2 and OMNI are aligned to GRCh37, whilst ADNI1 is aligned to GRCh36. For ANM, like ADNI1, samples were typed using the Illumina Human610-Quad BeadChip, but data had been moved to GRCh37 prior to acquisition.

### MR methodology

Inverse weighted variance (IVW) regresses exposure SNP-instrument associations with outcome SNP-instrument associations, weighted by the inversed variance of outcome SNP-instrument associations [81]. In IVW, the intercept is constrained to zero under the assumption that there is no horizontal pleiotropy. Odds ratios (OR) per 1 standard deviation were calculated to enable comparison to other exposures. Two robust methods, MR-Egger and weighted median [81], were used to generate alternative causal estimates. MR-Egger removes IVW’s intercept constraint with large deviations from zero at the intercept and between IVW and egger causal estimates providing evidence of horizontal pleiotropy [82]. Weighted median MR controls for bias in its causal estimate, even if up to 50% of the instruments are invalid, by ordering and weighting estimates by association strength and taking the estimate at the 50^th^ percentile [81]. Lastly, leave one out analysis was conducted to estimate the impact of individual SNPs and Cochran’s Q was calculated to test for between SNP heterogeneity.

### Protein heritability analysis

Average h^2^ across the proteins was 0.10 (see full results at https://alexhandy1.shinyapps.io/ad-genetic-overlap-web-results/), however, results were treated as indicative given the average standard error was 0.16 (including 8 proteins with h^2^ less than 0). There are also known issues of applying LDSR to samples less than 5000 (see https://github.com/bulik/ldsc/wiki/FAQ). Genetic correlation analysis was limited by the same lack of power with 63% of pairwise correlations not able to be calculated (“NA”), an average standard error of 2.3 and only 2 pairwise correlations with a nominal p-value less than 0.05.

### AD genetic data preparation

Following alignment of genetic QC with procedures applied to all participant GERAD1, ADNI and ANM data, 3,691,311 variants were available for analysis from the 5,218,413 overlapping variants across cohorts. PRSice removed a further 668,813 variants that did not overlap with the AD base data.

### Supplementary Tables

**Supplementary Table 1:** Summary characteristics of Sun et al participants selected randomly in two subcohorts from the INTERVAL study.

|  | Subcohort 1 (n=2481) | Subcohort 2 (n=820) |
| --- | --- | --- |
| Age (years and SD) | 43.6 (14.3) | 44.1 (14.2) |
| Sex (% male) | 51.6% | 49.5% |
| BMI (and SD) | 26.3 (4.8) | 26.6 (4.9) |
| Current smokers (%) | 8.6% | 8.5% |
| Current alcohol use (%) | 92.6% | 91.0% |

**Supplementary Table 2:**
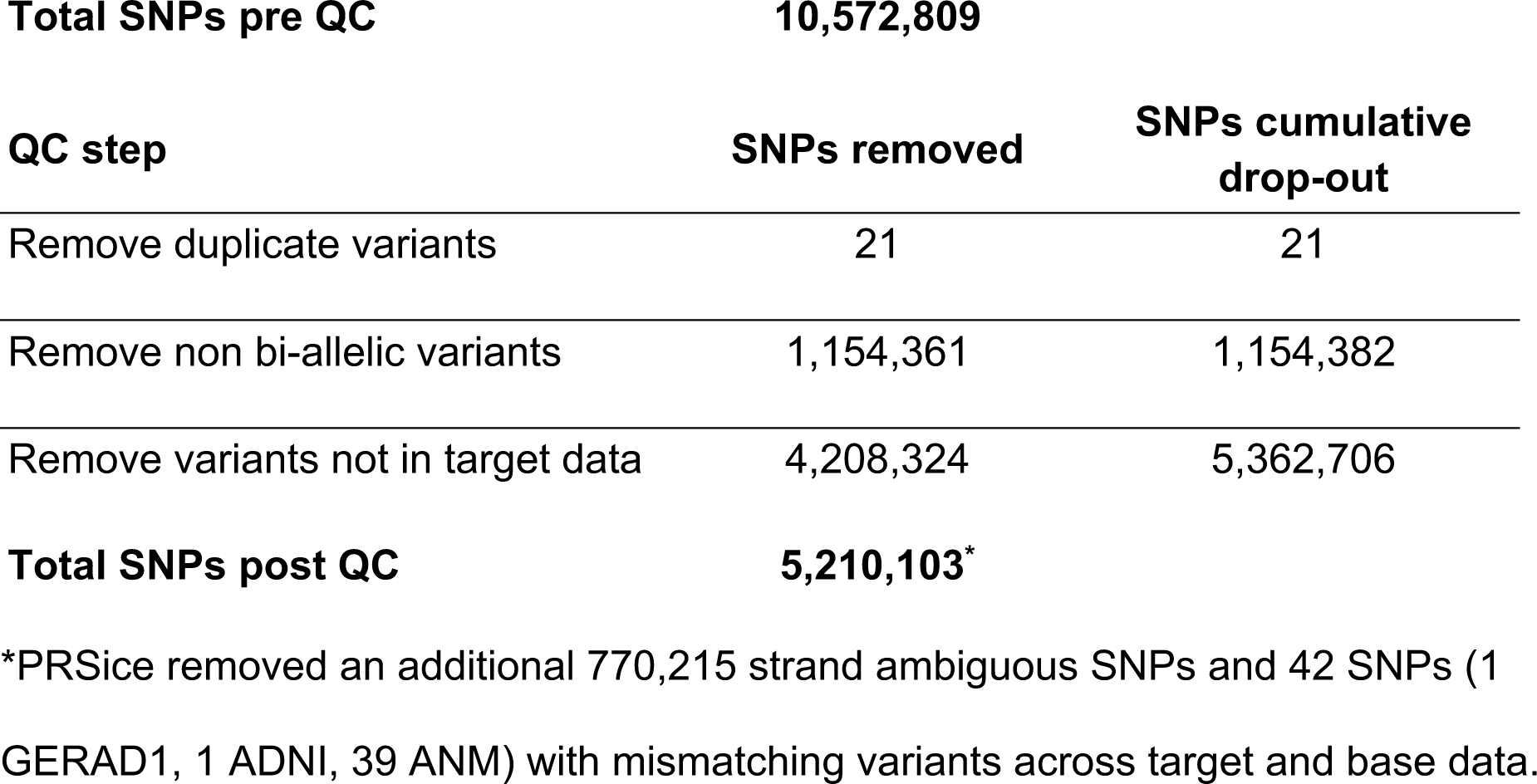
Total SNPs remaining for each protein in shortlist after QC.

